# Immunogenicity and Safety of the inactivated SARS-CoV-2 vaccine (BBIBP-CorV) in patients with malignancy

**DOI:** 10.1101/2021.09.02.21262760

**Authors:** Mona Ariamanesh, Pejman Porouhan, Babak PeyroShabany, Danial Fazilat-Panah, Mansoureh Dehghani, Maryam Nabavifard, Farbod Hatami, Mohammad Fereidouni, James S. Welsh, Seyed Alireza Javadinia

## Abstract

**Objective:** Patients with malignancy suffer from a compromised immune system due to either the effects of malignancies or treatments. Cancer patients are at higher risk of different infections particularly SARS-CoV2 and usually produce weaker response to vaccines. The aim of this study was to evaluate the safety and immunogenicity of the inactivated SARS-CoV-2 vaccine (Sinopharm, BBIBP-CorV) in patients with malignancy.

**Material and Method:** In total 364 patients with cancer (median age: 54 years old, F/M ratio: 217/147) who received two doses of Sinopharm vaccine were enrolled in this study. Vaccine related side effects was assessed by a questionnaire and the presence of SARS-CoV-2 anti-Spike protein (S) IgG and neutralizing antibody two months following vaccination were measured by immunological methods.

**Results:** Injection site pain and fever were the most common local and systemic side effects in vaccine receivers. Two months after the first dose, anti-S IgG and neutralizing antibody were detectable in 77.1% and 80.7% of all participants, respectively with an overall response to either or both measured in 86.9% of patients The rate of seroconversion was lower in older age, those with hematological malignancies and chemotherapy receivers.

**Conclusion:** The result of study confirmed the safety and short-term efficacy of Sinopharm inactivated vaccine (BBIBP-CorV) in patients with different type of malignancies.

## 1. Introduction

The outbreak of COVID-19 led to prompt development and production of vaccines in less than a year. To date, only healthy adults or those with underlying stable medical conditions were systematically assessed regarding the efficacy, immunogenicity, and safety of vaccines including Moderna, Oxford-AstraZeneca, Sputnik, and Sinopharm BBIBP-CorV in phases I, II, and III clinical trials. Given the extensive disease burden that the world has been facing, health control agencies across the world including the World Health Organization (WHO) and the United States Food and Drug Administration (FDA) of granted the emergency authorization for use of vaccines based on interim results of trials (1).

Multiple studies have demonstrated that patients with cancer are at increased risk for more severe disease and higher mortality from COVID-19 compared to healthy individuals (2-6). Also, recent studies have reported that, during the pandemic, newly diagnosed patients with cancer have presented with more advanced cancer stage associated with delays in cancer screening compared to a similar time frame before the pandemic (7). Moreover, many patients postponed treatment during the height of the pandemic due to fears of infection with SARS-CoV-2, resulting in worsening prognosis (8, 9). Therefore, establishing vaccine effectiveness and safety in patients with cancer to prioritize them in vaccination strategies is of crucial importance (10).

In Iran, the Food and Drug Administration approved the Sinopharm vaccine (BBIBP-CorV) with an emergency authorization in March 2021. National vaccination guidelines encouraged patients with cancer to receive the vaccine, although the decision to vaccinate patients undergoing active treatment was left to the attending oncologist. To our knowledge, there is no available information concerning COVID-19 vaccine BBIBP-CorV efficacy in patients with active cancer. The current ongoing study is the first aimed at assessing the immunogenicity and safety of the BBIBP-CorV vaccine in a cancer population.

## 2. Methods

### 2.1. Population of Study

All patients with cancer referred for vaccination to our cancer care network in the cities of Sabzevar, Neyshabur, and Babol were invited to participate in the study between March and June 2021. Patients with acute conditions, including infection and immune-related complications, were excluded. The protocol of the study was approved by the Ethics Committee of Sabzevar University of Medical Sciences (IR.MEDSAB.REC.1400.027) and a written informed consent form was obtained from the patients or the legal guardian.

### 2.2. Measurements

At baseline, the previous history of confirmed COVID-19 with real-time polymerase chain reaction (PCR) was assessed and blood samples were drawn to measure anti-SARS-CoV-2 Nucleocapsid (N) IgG [PISHTAZTEB DIAGNOSTICS, Tehran, Iran]. Subsequently, two doses of 0.5 mL Sinopharm β-propiolactone-inactivated, aluminum hydroxide-adjuvanted COVID-19 vaccine (BBIBP-CorV) were administered intramuscularly 28 days apart. Two months following vaccination, blood samples were drawn to analyze the presence of SARS-CoV-2 anti-Spike protein (S) IgG and neutralizing antibodies. To evaluate the vaccine immunogenicity, the level of SARS-CoV-2 Anti-Spike IgG, and SARS-CoV2 Anti RBD IgG were measured by two commercial ELISA kits [PISHTAZTEB DIAGNOSTICS, Tehran, Iran]. The sensitivity, specificity, and accuracy of both kits were 100% (95%CI 96.4-100), 99% (95%CI 94.9-99.9), and 99.5% (95%CI 97.4-99.9) respectively. According to the kits’ manual, a cut-off points of 8 μg/ml and 2.5 μg/ml were considered as positive response for the SARS-CoV-2 Anti-Spike IgG and SARS-CoV2-neutralizing antibody respectively.

Participants were followed for three months to evaluate short-term side-effects. Data regarding local and systemic side-effects were collected weekly via telephone or in-person using a questionnaire based on the Common Terminology Criteria for Adverse Events (CTCAE) version V. Furthermore, a hotline was established for the report of any serious acute side-effects. Diagram 1 shows the study protocol.

**Diagram 1:**
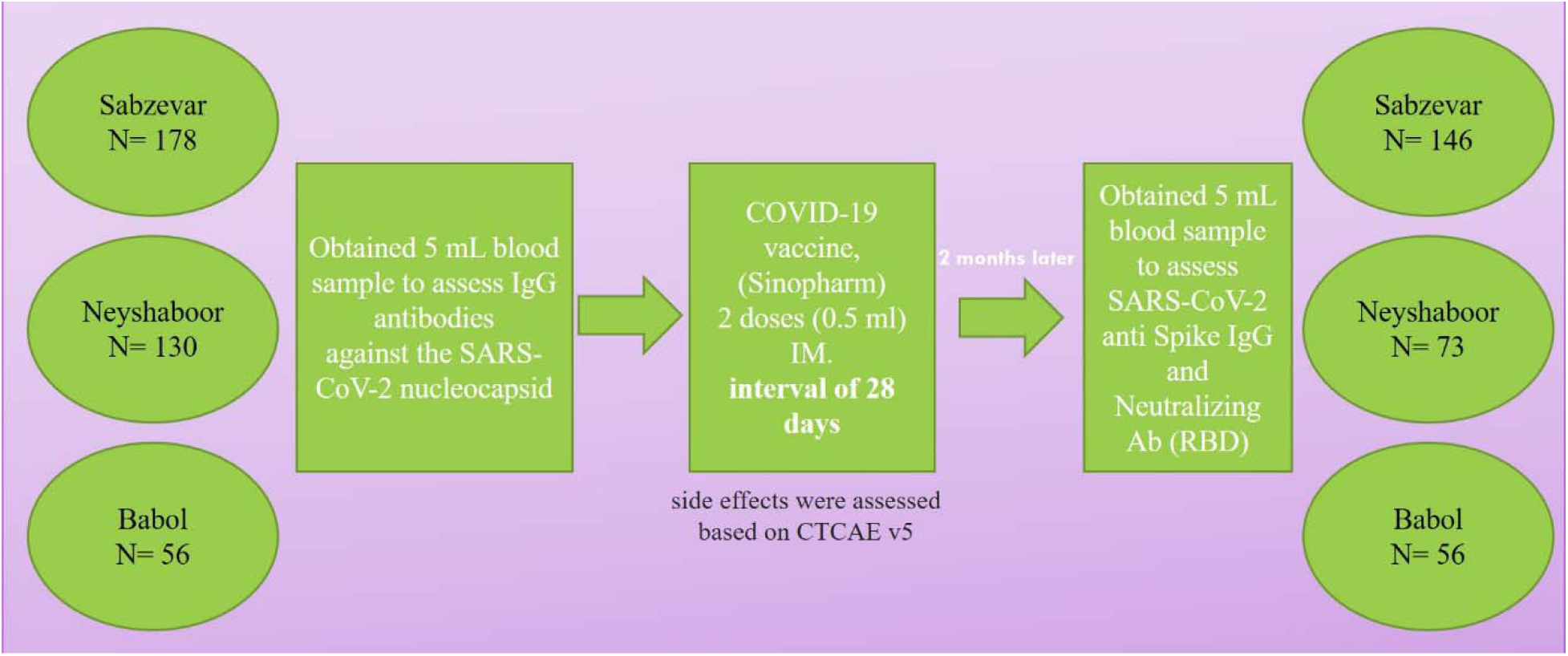
Study protocol.

### 2.2. Statistical Analysis

The primary outcome of this analysis was the proportion of patients with cancer developing positive serology for either the SARSCOV2 spike protein and/or COVID-19 neutralizing antibody two months following vaccination with the Sinopharm COVID-19 produced by the Beijing Bio-Institute of Biologic Products, a subsidiary of China National Biotec Group approved by WHO for Emergency use the Sinopharm BBIBP-CorV vaccine. Secondary outcomes include the proportion of patients testing positive for COVID-19 by PCR at one, two or three months following the first dose of vaccine as well as the proportion of patients reporting one or more adverse events during the three months following vaccination. Univariable and multivariable analyses explored the association of baseline covariates including prior PCR confirmed COVID-19 and those presenting with positive serology for SARSCoV2 IgG prior to vaccination. Continuous variables are summarized on the basis of means or medians while categorial variables are presented as proportions. Bivariate analysis for categories variables were assessed using the Pearson Chi-Square method with 2-sided tests for asymptotic significance. Multivariable analysis was based on logistic regression modeling using forward stepwise variable selection. Collinearity was assessed by estimation of variance inflation factor (VIF) of model covariates.

## 3. Results

### 3.1. Study Population

A total of 364 patients with cancer aged ≥18 years were evaluated in this prospective multi-center study. Average patient age was 54 years (range: 18-85). Participants were categorized into three age groups. Those aged 40-60 years were the most frequent group (n=194 people, 53.3%), followed by those aged >60 years (n=111 people, 30.5%), and those aged <40 years (n=59 people, 16.2%). Females and males comprised 59.6% and 40.4% of the study population, respectively.

Patients with both solid and hematological malignancies were included, with breast cancer being the most common type of malignancy among the participants (44%). Patients with other types of cancer included colorectal adenocarcinomas (14.6%), upper gastrointestinal cancers (8.8%), hematologic malignancies (6.6%), prostate adenocarcinomas (5.5%), head & neck squamous cell carcinomas (5.2%), gynecological cancers (2.8%), and brain gliomas (2.5%). The majority of our population had a progressive cancer stage III (56.3%), while there were 20.9% stage II, 11.5% stage IV, and 2.2% stage I of cancer. Of the total, 180 (49.4%) patients were on active treatment, of which 131 (72.7%) were receiving chemotherapy with or without radiotherapy, and 49 (27.3%) were receiving radiotherapy alone while 184 (5.5%) cases were follow up patients (Table 1).

**Table 1.** Patient characteristics, types of cancer, and treatments

|  | Frequency (%) |
| --- | --- |
| Age |  |
| <40 years | 59 (16.2) |
| 40-60 years | 194 (53.3) |
| >60 years | 111 (30.5) |
| Gender |  |
| Male | 147 (40.4) |
| Female | 217 (59.6) |
| Stage |  |
| I | 8 (2.2) |
| II | 76 (20.9) |
| III | 205 (56.3) |
| IV | 42 (11.5) |
| Cancer |  |
| Breast Cancer | 160 (44) |
| Prostate Cancer | 20 (4.9) |
| Upper GI Cancers | 32 (8.8) |
| Colorectal Cancer | 53 (14.6) |
| Brain Glioma | 9 (2.5) |
| Head & Neck Cancer | 19 (5.2) |
| Hematologic Malignancies | 24 (6.6) |
| Gynecological Cancers | 10 (2.7) |
| Others | 37 (10.2) |
| Treatment |  |
| Chemotherapy +/- RT | 131 (36) |
| Radiotherapy alone | 49 (13.5) |
| Follow up patients, including breast cancer<br>patients on endocrine therapy or<br>trastuzumab | 184 (50.5) |

Of the total population, 47 (12.9%) patients had a clinical history of COVID-19 prior to vaccination. However, based on the serological test of anti-N IgG, 23.4% of patients were seropositive at the time of vaccination. Two months following the first dose, anti-S IgG and neutralizing antibodies were detectable in 77.1% and 80.7% of all participants respectively with 86.9% of patients demonstrating immunity against COVID-19 when considering either anti-S or neutralizing antibodies. Patients were followed for three months post-vaccination. Four patients were infected with SARS-CoV-2 following the first dose with one of these four receiving the second dose of vaccine.

### 3.2. Immunologic Response

Overall, a post-vaccination positive response of IgG against the SARS-CoV-2 Spike protein or neutralizing antibody against RBD or both was observed in 239 (86.9%) patients. As shown in Table 2, The rate of seroconversion was higher in patients younger than 60 years (90.9%, 90%, and 79% in patients<40 years, 40-60 years, and >60 years; respectively, P=.042). This relationship with age was particularly evident for IgG response to the spike protein which was under 60% in those 60 years of age or older (P<.001). The rate of seroconversion was higher in patients with breast cancer (93.3%) and upper GI cancers (94.7%) and the lowest rate was for patients with hematologic malignancies (61.9%) which only 38.1% and 52.4% of the them were positive for anti spike protein and neutralizing antibodies respectively. Furthermore, any antibody response was significantly lower in patients receiving chemotherapy (83.5%) compared to over 97% of those receiving radiotherapy alone or endocrine therapy (P=.004). Similarly, response to SARS-CoV-2 Spike protein was positive in 70% in those receiving chemotherapy compared to 93% and 87% in those receiving radiation therapy alone or endocrine therapy, respectively. Of note, patients with a prior history of COVID-19 by PCR experienced a more robust response of the SARS-CoV-2 Spike protein than those without such history (P=.031) as did patients with positive SARS-CoV-2 IgG prior to vaccination (P=0.004).

**Table 2.** Serologic responses following SARS-CoV-2 vaccination by patient characteristics, type of cancer, and treatment.

|  | SARS-CoV-2 Spike Protein Positive (%) | COVID-19 Neutralizing Antibody Positive (%) | Either SARS-CoV-2 Spike Protein or Neutralizing Antibody Positive (%) |
| --- | --- | --- | --- |
| Age |  |  |  |
| <40 years | 37 (84.1) | 36 (81.8) | 40 (90.9) |
| 40-60 years | 127 (84.7) | 125 (83.3) | 135 (90) |
| >60 years | 48 (59.3) | 61 (75.3) | 64 (79) |
| P value | **<0.001** | 0.330 | **0.042** |
| Gender |  |  |  |
| Male | 83 (72.2) | 87 (75.5) | 95 (82.6) |
| Female | 129 (80.6) | 135 (84.4) | 144 (90) |
| P value | 0.100 | 0.70 | 0.073 |
| Stage |  |  |  |
| I | 6 (75) | 8 (100) | 8 (100) |
| II | 44 (78.6) | 44 (78.6) | 48 (85.7) |
| III | 126 (80.8) | 132 (84.6) | 139 (89.1) |
| IV | 20 (83.3) | 18 (75) | 95.8 (23) |
| P value | **0.010** | 0.053 | **0.009** |
| Cancer |  |  |  |
| Breast Cancer | 102 (85.7) | 104 (87.4) | 111 (93.3) |
| Prostate Cancer | 12 (75) | 11 (68.8) | 13 (81.3) |
| Upper GI Cancers | 18 (94.7) | 17 (89.5) | 18 (94.7) |
| Colorectal Cancer | 27 (65.9) | 31 (75.6) | 35 (85.4) |
| Brain Glioma | 6 (66.7) | 7 (77.8) | 7 (77.8) |
| Head & Neck Cancer | 11 (64.7) | 13 (76.5) | 13 (76.5) |
| Hematologic<br>Malignancies | 8 (38.1) | 11 (52.4) | 13 (61.9) |
| Gynecological<br>Cancers | 4 (80) | 4 (80) | 4 (80) |
| Others | 24 (85.7) | 24 (85.7) | 25 (89.3) |
| P value | **<0.001** | **0.021** | **0.010** |
| Treatment |  |  |  |
| Chemotherapy +/- RT | 68 (70.1) | 74 (76.3) | 81 (83.5) |
| Radiotherapy | 37 (92.5) | 37 (92.5) | 39 (97.5) |
| Follow-up | 41 (87.2) | 45 (95.7) | 46 (97.9) |
| P value | **0.008** | **0.001** | **0.004** |
| Previous COVID-19<br>PCR Report |  |  |  |
| Yes | 31 (88.6) | 32 (91.4) | 32 (91.4) |
| No | 191 (79.6) | 180 (75) | 180 (75) |
| P value | **0.031** | 0.208 | 0.396 |
| SARS-CoV-2 IgG<br>positive prior to<br>vaccination |  |  |  |
| Yes | 56 (88.9) | 57 (90.5) | 58 (92.1) |
| No | 166 (78.3) | 155 (73.1) | 181 (85.4) |
| P value | **0.004** | 0.061 | 0.167 |

In multivariable logistic regression analysis, only younger age and increasing stage were independently associated with combined immunity whereas younger age, upper GI cancer and positive pre-vaccination SARS-CoV-2 Ig were independently associated with SARS-CoV-2 Spike protein following vaccination.

Finally, five patients experienced confirmed breakthrough COVID-19 infections during the three months following vaccination, four of which occurred during the first month. None of these patients had prior COVID-19 or positive SARS-CoV-2 Ig prior to vaccination. Three were being followed without any cancer treatment and two had stage III disease. None of our patients died nor were admitted to the hospital during the study period.

### 3.3 Adverse Events Following Vaccination

local side-effects included mild, moderate, and severe pain was recorded in 15.4%, 9.3%, and 2.2%, of vaccine receivers respectively. Other local side-effects were injection site redness (2.2%), swelling (1.1%), and itching (0.7%). Fever (31.6%) was the most common systemic side-effect observed while chills (11.5%), fatigue (21.6%), anorexia (13.5%), nausea (10.4%), vomiting (2.9%), myalgia (19.4%), and diarrhea (2.9%) were less common. Detailed systemic side-effects are presented in Tables 3. Injection site pain and generalized myalgias were somewhat more common in younger patients and females but occurred in all age groups and both genders. High grade fever (temperature>39°C) was more commonly observed in females following vaccination (P=0.10). Injection site pain and swelling was more common in those with positive SARS-CoV-2 IgG prior to vaccination than those that were negative. Headache and myalgias were also more commonly reported in those with positive SARS-CoV-2 IgG prior to vaccination than among those who were negative.

**Table 3.**
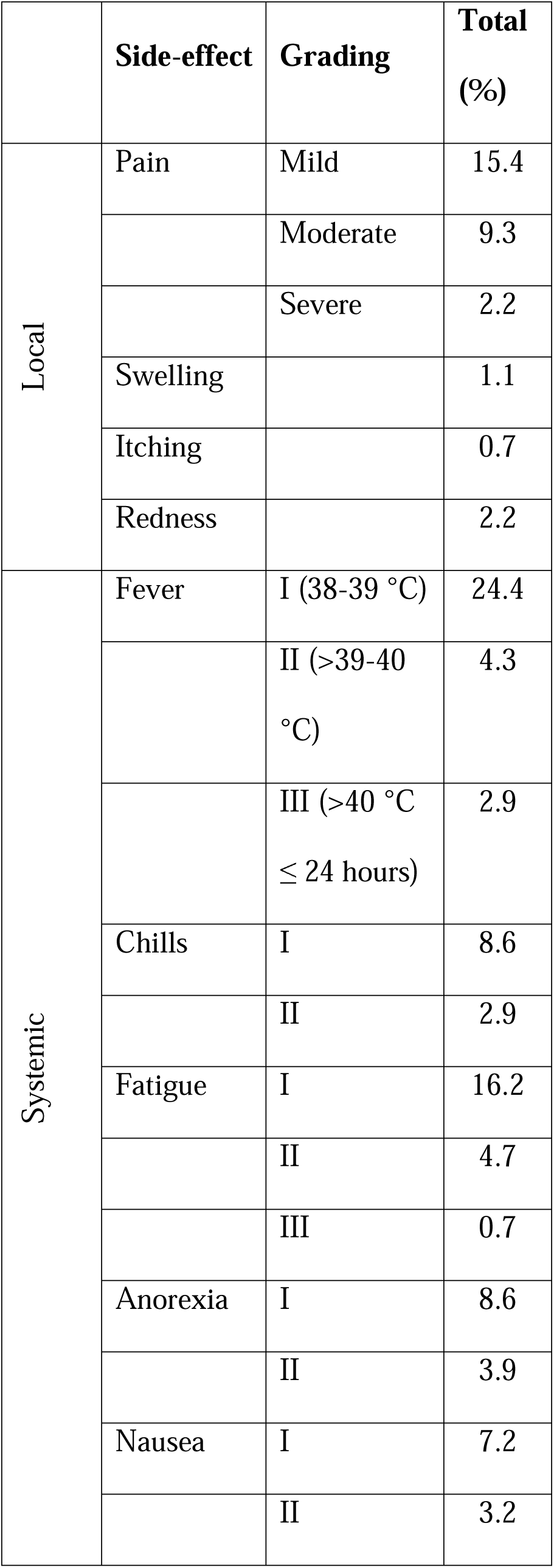
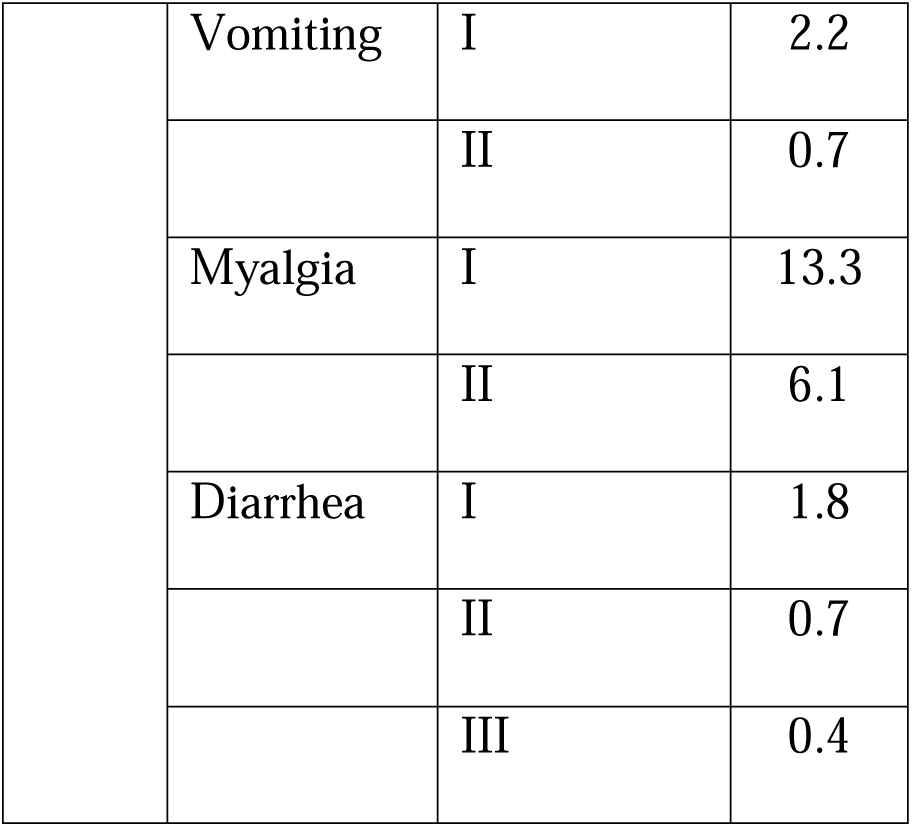
Local and systemic side-effects related to vaccine

## 4. Discussion

It is well-known that patients with active cancer might experience more adverse outcomes if they contract COVID-19. Early data from China showed the cancer population is at a 3.5-fold heightened risk of requiring mechanical ventilation and admission to an intensive care unit compared to the general population (11). Another investigation from the United Kingdom found a remarkable mortality rate of 28% due to COVID-19 in the cancer population (2). These patients are more susceptible to severe COVID-19 for two reasons; first, the nature of malignancy itself that induces immunosuppression and disturbances in metabolism, and second, anti-cancer treatments including cytotoxic chemotherapy and immunomodulatory agents which compromise appropriate response by memory B cells and antigen-specific T cells (12). Consistently, hematologic malignancies are associated with significantly higher mortality rates from COVID-19 compared with other cancers. Also, seroprevalence studies revealed that patients with cancer develop a lower level of IgG antibodies against SARS-CoV-2 following infection (72.5% seropositive) (13).

Despite a lack of adequate knowledge for use of SARS-CoV-2 vaccines in cancer patients, emergency authorization was granted by many agencies and health ministries around the world based on a prediction that benefits will overshadow possible detriments, coupled with past experience indicating advantages of influenza vaccination in cancer patients.

The BBIBP-CorV vaccine is an inactivated vaccine manufactured by Sinopharm Inc. and was approved by the China National Medical Products Administration on December 31, 2020, for use in adults aged ≥18 years. A Phase III clinical trial from Bahrain confirmed effectiveness of 78.1% in healthy adults (14). The most common local and systemic side-effects were pain at the injection site (18.8%) and headache (12.6%), respectively. Similar to our present results, fever was the most frequent systemic side-effect in phase I and II trials (15, 16). More recently, Li et al showed that inactivated SARS-CoV-2 vaccines are effective against Delta variant infection as well (17).

A recent report evaluated the safety of the Pfizer-BioNTech mRNA-based vaccine in oncology patients and showed no significant difference in side-effects between the healthy and cancer groups except for myalgia (34%) being the most common systemic side-effect, which was notably more common in the cancer group. Similar to our results, pain at the injection site (63%) was the most common local side-effect (18). Anecdotally, some physicians have been concerned that SARS-CoV-2 vaccination of any sort during a course of radiation therapy could cause side-effects that might cause their patients to miss a session or two. This concern was not confirmed in our study as no patients missed any radiotherapy sessions due to local or systemic side effects of vaccination.

Safety concerns for cancer patients were further addressed by another article following the administration of two shots of Pfizer vaccine with an interval of 21 days and the approximate follow-up period of three months. The study also evaluated the immunogenicity of the vaccine by measuring anti-S IgG and found seroconversion in 95%, 38%, and 18% after the first dose and 100%, 95%, and 60% after the second dose in healthy adults and those with solid cancers, and hematologic malignancies, respectively (19). These data are consistent with our findings of appropriate immunological responses in 86.9% of all cancer patients but only 61.9% of those with hematologic malignancies using the Sinopharm inactivated vaccine (BBIBP-CorV). Finally, our findings that any antibody response was observed in 83.5% of patients actively receiving chemotherapy was encouraging, and our findings that over 97% of those undergoing endocrine therapy or radiotherapy alone responded well to the vaccinations was quite reassuring.

However, in our present study, at only 61.9% the overall response to COVID-19 inactivated vaccine was suboptimal and significantly lower in patients suffering hematological malignancies than other types of cancers. Curiously, only 38.1% of our hematologic malignancy population developed anti-spike protein antibodies. Similarly, in a preprint manuscript Agha et al (2021) reported that under half of the patients with blood cancers did not produce detectable antibodies to the SARS-CoV-2 spike protein following two doses of the COVID-19 mRNA vaccines (20). In their report, only 23% of patients with B-cell CLL developed detectable antibodies. Recently, Ollila et al reported consistent results where they showed that only 39.3% of patients with hematologic malignancies demonstrated post COVID-19 vaccination seroconversion (21). Importantly, they found that the rate of seroconversion was significantly influenced by a recent treatment with B-cell/plasma cell-depleting monoclonal antibodies. Additionally they observed that patients with active malignant disease and those with a briefer time interval between vaccination and their last chemotherapy session had lower seroconversion rates (21). A recent paper by Massarweh et al showed that seroconversion was adequate in cancer patients following two doses of vaccination using BNT 162b2, which is another mRNA vaccine. That study only evaluated rates of anti-Spike antibodies whereas our study investigated rates of anti-Spike and neutralizing antibodies. However, their study also examined specific titers of anti-Spike IgG levels and discovered that median titers were somewhat lower in cancer patients compared to healthy controls. Multivariable analysis showed that the only variable statistically significantly associated with lower IgG titer was treatment with chemotherapy plus immunotherapy (22). Although the evidence are lacking, it seems comorbid cancer and treatment with systemic therapy, including chemotherapy, may influence the immune response to SARS-CoV-2 and potentially COVID-19 vaccination (23).

In the current study, a fairly large group of patients with various malignancies were carefully evaluated for vaccine-related side effects and their humoral response against vaccine was measured by two different methods. There are some limitations. For example, although this study assessed the short-term serologic and clinical efficacy of BBIBP-CorV vaccine, longer follow up is essential to confirm the long-term effects, need for further boost dose, and efficacy against newer variants of SARS-CoV-2.

## 5. Conclusion

In conclusion, the results of this study confirmed the safety and short-term efficacy of Sinopharm inactivated vaccine (BBIBP-CorV) in patients with malignancy, although the rate of seropositivity was lower in those of older age, those suffering from hematologic malignancies, or patients actively receiving chemotherapy. The Sinopharm inactivated vaccine appears to be safe and very effective in cancer patients receiving only radiation therapy or hormonal therapy but has lower rates of seroconversion among those actively undergoing cytotoxic chemotherapy. At only 61.9%, seroconversion rates were lowest among those with hematologic malignancies using the Sinopharm inactivated vaccine. Additional measures may be needed to prevent COVID-19 in cancer patients undergoing chemotherapy and those with hematologic malignancies. Further studies over longer periods of time are needed to fully evaluate the trend of humoral response and long-term efficacy of inactivated vaccines in cancer patients, particularly against newer variants of SARS-CoV-2.

## Data Availability

All data generated and analyzed during this study can be accessed through direct communication with the corresponding author and the agreement of all research team members.

## Acknowledgements

This research was funded by Sabzevar University of Medical Sciences (grant number 400004) and Neyshabur University of Medical Sciences. Authors would like to thank all patients who participate in the project. Also, a special thanks to Ms. Batol Keykhosravi and Ms. Azam Akbari Yazdi for their great helps. Moreover, authors truly appreciate all efforts have been done by Dr. Gary H. Lyman for data analyzing and critical review of the article.

## Notes

**Conflict of interest statement** The authors declare that there is no conflict of interest to be reported.

### Competing Interest Statement

The authors have declared no competing interest.

### Author Declarations

The protocol of the study was approved by the Ethics Committee of Sabzevar University of Medical Sciences (IR.MEDSAB.REC.1400.027) and a written informed consent form was obtained from the patients or the legal guardian.

